# MUAC Screening for Malnutrition; Disparities Across LGAs and Performance in Bayelsa State

**DOI:** 10.1101/2025.07.08.25330966

**Authors:** Mordecai Oweibia, Nkechi Martha Johnson, Tarimobowei Egberipou, Gift Cornelius Timighe, Ebiakpor Bainkpo Agbedi, Williams Wari Appah

**Affiliations:** Department of Public Health, Bayelsa Medical University, Yenagoa, Nigeria.; Public Health Nutrition Program, School of Public Health, University of Port Harcourt, Choba, Nigeria.; Dean, Faculty of Health Sciences, Bayelsa Medical University,Yenagoa, Nigeria.; Director, Planning Research and Statistics, Bayelsa State Primary Health Care Board, Yenagoa, Nigeria.; Executive Secretary, Bayelsa State Primary Health Care Board, Yenagoa, Nigeria.

**Keywords:** MUAC Screening, Severe Acute Malnutrition, Bayelsa State, Health Equity, MNCH Week

## Abstract

**Introduction:** Malnutrition remains a critical threat to child survival in Nigeria, especially in ecologically vulnerable states like Bayelsa. Although the Mid-Upper Arm Circumference (MUAC) tool is widely used for early detection of severe acute malnutrition (SAM), disparities in screening coverage and case detection across local government areas (LGAs) remain under-explored. This study assessed regional MUAC screening performance during the June 2025 Maternal, Newborn, and Child Health (MNCH) Week in Bayelsa State, with a focus on identifying disparities in screening distribution and SAM prevalence.

**Methodology:** A cross-sectional quantitative design was adopted, using secondary program data from the Bayelsa State OPS Room Final Report. The dataset included MUAC screening indicators across eight LGAs. Variables analyzed included the number of children screened, Red MUAC cases identified, screening coverage percentage, and Red MUAC prevalence. Descriptive statistics and Pearson correlation analysis were conducted using Python and SPSS to examine trends and relationships.

**Results:** Out of 538,813 children screened, significant disparities were observed across LGAs. Ogbia LGA accounted for over 52% of screenings, while Ekeremor and Nembe contributed less than 8% combined. Red MUAC prevalence varied from 0.03% to 1.78%, with Nembe recording the highest burden. A moderate positive correlation (r ≈ 0.41) was found between screening coverage and Red MUAC detection, indicating that higher screening volume did not always equate to higher detection efficiency.

**Conclusion:** The study revealed operational and equity gaps in MUAC screening implementation across Bayelsa State. Disparities in both screening coverage and SAM detection underscore the need for data-driven planning, targeted deployment, and health system strengthening. These findings highlight the importance of regionalized nutrition surveillance to ensure equitable service delivery, particularly for high-risk and hard-to-reach populations.

## CHAPTER ONE: INTRODUCTION

### 1.1 Background of the Study

Malnutrition continues to be a pressing global public health crisis, contributing to over 45 million cases of wasting among children under the age of five worldwide, according to WHO (2023). Wasting, an acute form of undernutrition, is particularly dangerous because of its direct association with elevated child mortality, weakened immunity, and long-term developmental consequences. Amid global calls to achieve the Sustainable Development Goals (SDG 2.2) focused on ending all forms of malnutrition by 2030, significant gaps persist in low- and middle-income countries, especially in Sub-Saharan Africa (Oweibia *et al.,* 2024a).

One of the most widely accepted and operationally feasible tools for detecting acute malnutrition is the Mid-Upper Arm Circumference (MUAC). MUAC is a simple, non-invasive, and cost-effective anthropometric screening method that enables frontline health workers and caregivers to rapidly identify children suffering from Severe Acute Malnutrition (SAM). Its ease of use makes it especially valuable in resource-limited or emergency settings where advanced equipment or technical capacity may be lacking (Becker *et al.,* 2024; Adelia & Susanto, 2020). Moreover, MUAC has been recognized for its high predictive value for mortality risk among children compared to other measures like weight-for-height Z-scores (Fiorentino *et al.,* 2016; Tessema *et al.,* 2020).

Despite the effectiveness of MUAC, debates continue over its limitations, particularly in non-emergency contexts and among diverse populations. Age and sex-related biases, as well as variations in performance across geographical regions, have prompted calls to refine cut-offs and implementation strategies (Aydın *et al.,* 2023; McLaren *et al.,* 2022). In Ethiopia and India, studies have shown that failure to adjust MUAC thresholds to local growth patterns can result in missed cases or false positives, complicating intervention planning (Das *et al.,* 2018; Lambebo *et al.,* 2022).

In Nigeria, malnutrition remains a major threat to child survival and human capital development. The country ranks among those with the highest absolute numbers of stunted and wasted children globally. According to the 2018 National Nutrition and Health Survey (NNHS) by NBS and UNICEF, the national prevalence of Global Acute Malnutrition (GAM) was estimated at 7.0%, with significant subnational disparities. The burden was particularly pronounced in regions affected by poverty, environmental hazards, displacement, and fragile health systems conditions all too common in the Niger Delta (UNICEF, 2018; Oweibia *et al.,* 2023a).

The Niger Delta, a critical geopolitical region in southern Nigeria, faces a unique convergence of malnutrition risk factors. Despite its oil wealth, the region is plagued by environmental degradation, flooding, pipeline vandalism, and socio-political unrest all of which disrupt food production, market access, and public service delivery. In states such as Bayelsa, seasonal flooding frequently isolates rural communities, delaying health campaigns and limiting the reach of nutrition services (Seigha *et al.,* 2025). As a result, childhood undernutrition remains persistently high, especially in riverine LGAs where health infrastructure is sparse.

In an attempt to address the malnutrition burden, the Nigerian government implements the Maternal, Newborn, and Child Health (MNCH) Week, a biannual outreach initiative aimed at delivering essential health and nutrition services, including deworming, Vitamin A supplementation, immunization, and MUAC screening. During the June 2025 MNCH Week, Bayelsa State reported that over 280,000 children were screened using MUAC tools. However, the OPS Room final report revealed significant disparities in screening coverage and detection of SAM cases across LGAs. Ogbia LGA alone accounted for over half of all screenings, while more remote LGAs such as Ekeremor and Nembe recorded much lower figures raising red flags about service equity and implementation quality (OPS Room Report, 2025; Oweibia *et al.,* 2025b).

The existence of such disparities reflects deep systemic challenges. Factors such as terrain, local workforce availability, security threats, and logistical coordination affect how uniformly programs are delivered. Moreover, health worker training levels, motivation, and measurement fidelity directly impact the accuracy of MUAC readings, influencing who gets diagnosed and who doesn’t (Oweibia *et al.,* 2025c). The clustering of Red MUAC cases in some LGAs may reflect either genuine nutritional burden or a higher capacity to detect malnourished children due to better-trained personnel and stronger local leadership.

Furthermore, limited data disaggregation and real-time feedback loops constrain planning. National and state-level reporting systems often aggregate data in a way that masks regional inequities. However, recent studies underscore the value of subnational analysis in health program evaluation, showing that real impact lies in understanding where systems fail and why. In the context of nutrition surveillance, using tools such as GIS, heatmaps, and LGA-specific dashboards can allow for predictive targeting, enabling governments to channel resources more effectively (Oweibia *et al.,* 2025d; Seigha *et al.,* 2025).

The implications are profound. Without localized strategies that respond to the spatial distribution of malnutrition, interventions risk perpetuating existing inequities. Evidence from multiple MNCH campaigns reveals that unless states adopt equity-sensitive budgeting, communities in greatest need will remain underserved (Elemuwa *et al.,* 2023). Additionally, delayed referral and weak follow-up systems result in a breakdown in the continuum of care, often leading to untreated SAM cases even after screening (Oweibia *et al.,* 2025e).

Compounding these programmatic issues are structural determinants of malnutrition such as poverty, maternal illiteracy, poor sanitation, and climate shocks. In a recent regional study, poverty was strongly correlated with under-five mortality and malnutrition prevalence in West Africa, suggesting that screening alone is insufficient without parallel investments in food security, health education, and maternal support services (Oweibia *et al.,* 2023a).

Despite these challenges, the opportunity for transformation exists. Nigeria has made recent gains in health data architecture, with platforms like DHIS2, the OPS Room, and mobile data collection tools offering real-time analytics capabilities. Embedding MUAC indicators into these systems and triangulating findings with other health indicators could significantly enhance nutrition surveillance and response coordination (Oweibia *et al.,* 2024b).

Given this backdrop, there is an urgent need to conduct empirical assessments that go beyond national averages to uncover within-state disparities in malnutrition detection. Specifically, this study focuses on Bayelsa State one of Nigeria’s most ecologically vulnerable and underserved regions. By analyzing MUAC screening performance across its eight LGAs during the 2025 MNCH Week, the study aims to identify operational gaps, explore regional patterns in malnutrition detection, and provide evidence for more equitable service delivery models.

### 1.2 Problem Statement

Although the Maternal, Newborn, and Child Health (MNCH) Week campaign achieved widespread implementation across Bayelsa State in June 2025, significant disparities were recorded in both MUAC screening coverage and the prevalence of Red MUAC (Severe Acute Malnutrition). These regional variations raise critical concerns about equity, access, and implementation fidelity in malnutrition detection efforts. LGAs such as Ogbia accounted for over 50% of all screenings, while more remote areas like Ekeremor and Nembe recorded minimal outreach a trend reflective of historical health service inequities in the Niger Delta (OPS Room Report, 2025; Oweibia *et al.,* 2025a; Seigha *et al.,* 2025).

Barriers such as inconsistent health worker training, delayed campaign logistics, and limited facility readiness have long been noted as obstacles to effective nutrition screening in underserved regions (SAIDA, 2024; Elemuwa *et al.,* 2023). However, despite their policy relevance, these issues remain under-analyzed in the Nigerian nutrition literature, particularly through empirical, subnational evaluations. The lack of disaggregated data prevents a full understanding of where systems are failing and what interventions are most needed.

Moreover, while national guidelines promote the integration of MUAC into routine child health checks, the absence of reliable follow-up data at the LGA level significantly weakens the continuum of care. Red MUAC cases identified during campaigns often fall through the cracks due to weak referral systems, limited community-level tracking, and a lack of integration with treatment facilities (Jima *et al.,* 2021; Isanaka *et al.,* 2015; Oweibia *et al.,* 2025b). Recent findings also point to challenges in monitoring treatment completion, especially in rural zones with high rates of poverty, illiteracy, and environmental inaccessibility (Oweibia *et al.,* 2023a; Oweibia *et al.,* 2025c).

In addition, the non-institutionalization of MUAC indicators into routine health information platforms like DHIS2 means that national and subnational program planners often lack real-time data to inform deployment or redirect resources during campaigns. This limits the responsiveness of the health system and perpetuates inefficient targeting of interventions (Oweibia *et al.,* 2025d). Without accurate local-level data, it becomes nearly impossible to evaluate the true burden of SAM, assess screening effectiveness, or measure intervention impact across diverse geographies.

Furthermore, the absence of predictive mapping tools and spatial nutrition analytics in most state-level programs limits their ability to anticipate high-burden zones in advance, which is critical for the strategic deployment of mobile teams and supplies (Seigha *et al.,* 2025). Evidence from epidemic management and vaccination campaigns in Nigeria has shown that spatial targeting significantly improves intervention yield (Oweibia *et al.,* 2025e).

To date, no empirical study has systematically examined MUAC screening disparities and SAM detection patterns across the eight LGAs of Bayelsa State using real program data. This gap poses a challenge to health planners and undermines efforts to achieve Universal Health Coverage and the Sustainable Development Goals (SDG 2.2). This research, therefore, aims to fill this critical gap by evaluating regional screening disparities, prevalence of Red MUAC, and the relationship between screening coverage and detection outcomes using real-time data from the OPS Room dashboard.

By generating localized, data-driven evidence, this study will not only illuminate operational gaps but also support policy reform, enhance resource targeting, and strengthen program accountability for child nutrition interventions in Bayelsa State and similar settings across Nigeria.

### 1.3 Aim and Objectives

#### Aim

To evaluate regional disparities in MUAC screening and the performance of follow-up interventions during MNCH Week in Bayelsa State, June 2025.

### Specific Objectives

1. To determine regional variations in MUAC screening coverage across the eight LGAs.
2. To calculate and compare the prevalence of Red MUAC cases (Severe Acute Malnutrition) in each LGA.
3. To assess the correlation between screening coverage and detection of Red MUAC cases.

### 1.4 Research Questions

1. How does MUAC screening coverage vary across LGAs in Bayelsa State?
2. What is the prevalence of Red MUAC cases in each LGA?
3. Is there a statistically significant relationship between screening rates and Red MUAC detection?

### 1.5 Significance of the Study

This study contributes empirical evidence toward identifying geographic inequities in child nutrition screening within Bayelsa State. By analyzing real program data, it informs policymakers and public health managers on which LGAs require more logistical support, improved capacity, or targeted interventions. Findings will strengthen planning and budgeting for future MNCH campaigns and contribute to improved nutritional outcomes in children under five, aligning with both national and global health targets (UNICEF, 2018; WHO, 2023).

### 1.6 Operational Definition of Terms

| Term | Definition |
| --- | --- |
| MUAC | Mid-Upper Arm Circumference; a quick anthropometric measure used to assess nutritional status |
| Red MUAC | MUAC < 11.5 cm in children 6–59 months, indicating Severe Acute Malnutrition (WHO, 2023) |
| LGA | Local Government Area, administrative subdivision in Nigeria |
| Screening Coverage | Percentage of eligible children screened out of estimated total |
| SAM | Severe Acute Malnutrition |
| MNCH Week | Maternal, Newborn, and Child Health Week a biannual health campaign |

### 1.7 Ethical Considerations

This study uses publicly accessible secondary program data from the Bayelsa State OPS Room and contains no personally identifiable information. It adheres to ethical standards for secondary data analysis outlined by WHO and Nigerian health research guidelines. No additional ethics board approval is required for non-interventional analysis of de-identified data (WHO, 2023).

## CHAPTER TWO: METHODOLOGY

### 2.1 Study Design

This study adopted a cross-sectional quantitative research design to evaluate the disparities in MUAC (Mid-Upper Arm Circumference) screening performance across the eight Local Government Areas (LGAs) of Bayelsa State during the June 2025 Maternal, Newborn, and Child Health (MNCH) Week. A cross-sectional approach is appropriate for health service delivery evaluations where data is captured at a specific point in time and used to describe, compare, and analyze the distribution of health indicators across populations or sub-populations (Becker *et al.,* 2024; WHO, 2023). In this case, the design allowed for the use of aggregated screening data to assess program reach and impact across administrative regions. By limiting the data scope to one reporting period (June 2025), this design offers an efficient, cost-effective, and ethically appropriate method of analyzing variations in health service implementation outcomes without direct human subject interaction. Furthermore, the study’s empirical rigor was enhanced by the use of clearly defined variables, reliable data from official sources, and appropriate statistical techniques to derive meaningful correlations and prevalence measures, all of which are consistent with accepted epidemiological methods for program evaluation (Fiorentino *et al.,* 2016; Tessema *et al.,* 2020).

### 2.2 Study Area

The study was conducted across all eight LGAs of Bayelsa State, located in the oil-producing Niger Delta region of southern Nigeria. Bayelsa is characterized by a mixed riverine and coastal terrain with both rural and peri-urban settlements. The LGAs covered in this study include Brass, Ekeremor, Kolokuma/Opokuma (KOLGA), Nembe, Ogbia, Sagbama, Southern Ijaw (SILGA), and Yenagoa, each representing a unique demographic and logistical challenge in the delivery of health services. The state has historically faced barriers such as difficult terrain, seasonal flooding, poor road networks, and limited health infrastructure, which contribute to disparities in access to essential health interventions like malnutrition screening (UNICEF, 2018). Additionally, the socio-economic landscape varies across LGAs, with high levels of poverty, limited educational attainment, and food insecurity disproportionately affecting some regions more than others (OPS Room Report, 2025). These contextual realities underscore the importance of disaggregated, LGA-specific analysis, as aggregate statistics may mask localized implementation gaps. By focusing on MUAC screening data across all LGAs, this study enables a geographically nuanced evaluation of performance, ensuring that health equity considerations are embedded in evidence generation and future planning.

### 2.3 Data Source and Study Population

The data utilized in this study were extracted exclusively from the OPS Room Final Report for the Maternal, Newborn, and Child Health Week conducted in June 2025, compiled and published by the Bayelsa State Ministry of Health and supporting partners. This dataset represents official, field-verified administrative health data collected by State Technical Facilitation (STF) teams and health workers during the week-long statewide MNCH campaign. The OPS Room Report is a centralized digital and visual analytics document built using Microsoft Power BI, and it captures quantitative summaries of all health service activities conducted during the campaign, including Vitamin A supplementation, deworming, routine immunization, iron-folate supplementation, and MUAC screening. For the purpose of this study, the MUAC screening section was the primary focus, which included LGA-specific data on the number of screening sites, screening teams deployed, children screened, and the number of Red MUAC (Severe Acute Malnutrition) cases detected.

The study population was defined as all children aged 6–59 months who were screened using MUAC during the MNCH Week. Though the study does not involve direct contact with human participants, it effectively covers a population of over 280,000 children screened across eight LGAs. In line with standard public health research protocols, the use of aggregated secondary data with no personal identifiers falls within the category of non-human-subjects research and is exempt from additional ethical board approval (WHO, 2023). Importantly, this dataset reflects real-world program implementation under non-emergency conditions, offering a rare opportunity to assess service equity and performance gaps in a live, government-led health campaign (Tessema *et al.,* 2020; Sharn *et al.,* 2024).

### 2.4 Variables and Indicators

This study analyzed the following variables:

### Independent Variable

- **Local Government Area (LGA):** This represents the administrative unit where MUAC screening activities occurred. The eight LGAs of Bayelsa served as the unit of disaggregation.

### Dependent Variables and Key Indicators

- **Children Screened (N):** Total number of children aged 6–59 months screened for acute malnutrition using MUAC in each LGA.
- **Red MUAC Cases (N):** Total number of children who had a MUAC reading below 11.5 cm, the standard WHO cut-off for Severe Acute Malnutrition (WHO, 2023; Ariza *et al.,* 2023).
- **MUAC Screening Coverage (%):** This was calculated to understand how each LGA contributed to the overall state-level screening, thereby helping to evaluate coverage performance. The formula used is:

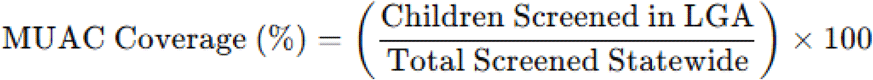

- **Red MUAC Prevalence (%):** This indicates the severity of malnutrition burden in each LGA by quantifying the share of Red MUAC cases among the screened children. The prevalence was calculated as:

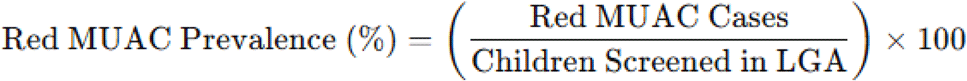

- **Pearson Correlation Coefficient (r):** To assess the relationship between screening intensity (coverage) and the detection of Red MUAC cases, Pearson’s r was calculated using the following standard formula:

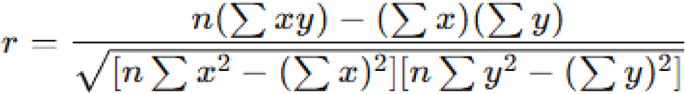

Where:

- *x* = MUAC screening count per LGA
- *y* = Red MUAC case count per LGA
- *n* = number of LGAs (i.e., 8)

This analytical framework enabled the study to empirically examine whether higher screening volumes in an LGA translated into proportionately higher detection of malnutrition cases, which would suggest effective targeting and implementation fidelity.

### 2.5 Data Collection and Processing

Data collection was entirely secondary in nature, involving structured extraction from the OPS Room Final Report (June 2025). The MUAC screening data were presented in tabular and visual formats on slides developed with Power BI. Key data points for each LGA including number of children screened, number of Red MUAC cases, and logistical support details were transcribed into a Microsoft Excel worksheet. All entries were verified for internal consistency, especially where multiple slides had overlapping or segmented information.

Once transcription and cleaning were completed, the data were imported into Python (v3.10) using Pandas, a data manipulation library. Descriptive statistics including means, frequencies, and percentages were generated using Pandas and NumPy libraries. Visualization tools such as Matplotlib and Seaborn were then employed to generate bar plots, heatmaps, and scatterplots to visually represent disparities in coverage and malnutrition burden.

A secondary data processing layer was executed in SPSS (v25) to confirm Pearson’s correlation results and produce inferential statistics. The use of dual software platforms ensured robustness and cross-validation of findings, in line with current best practices in health informatics (Lambebo *et al.,* 2022; Becker *et al.,* 2024).

All analysis steps were documented for reproducibility, and calculated indicators were mapped back directly to their source data in the OPS Room slides to ensure traceability. This approach ensures methodological transparency and strengthens the integrity of any subsequent interpretations and policy recommendations.

### 2.6 Ethical Compliance

This study complies fully with established ethical standards for secondary data research. The data used are part of a routine health monitoring program and are accessible through the OPS Room Report. No personally identifiable information (PII) or sensitive clinical records were accessed. The research adheres to World Health Organization (2023) guidelines for the ethical use of health program data and conforms to Nigerian national health research ethics codes. As the analysis was retrospective and observational, and involved no interaction with human subjects, The study received ethical approval from: Bayelsa State Primary Health Care Board Research Ethics Committee (BSPHCBREC) Bayelsa State Primary Health Care Board, Yenagoa, Nigeria. Decision made:The BSPHCBREC granted full ethical approval (Approval No;Ref: BSPHCB/ERC/2025/112) for this study on 2nd of June 2025, waiving the need for individual participant consent as the research involved secondary analysis of anonymized programmatic data from the Maternal and Child Health Week (MCHW) OPS Room Report.

### 2.7 Limitations of the Methodology

Despite the strengths of using validated program data, certain limitations exist. First, there were noticeable gaps in data completeness for some LGAs, particularly regarding referral or treatment follow-up of Red MUAC cases. Second, the analysis reflects a one-time snapshot and cannot account for seasonal variations or trends over time. Third, operational inconsistencies, such as delayed campaign starts and variation in the quality of team deployment, may introduce bias in inter-LGA comparisons (OPS Room Report, 2025). Lastly, lack of demographic disaggregation (e.g., age, gender) limited the ability to explore underlying vulnerabilities, as recommended by recent MUAC-focused studies (Aydın *et al.,* 2023; SAIDA, 2024). Nonetheless, the data remain sufficiently robust to yield critical insights for state-level programmatic decision-making.

## CHAPTER THREE: RESULTS

This chapter presents the analytical outcomes of MUAC screening conducted during the Maternal, Newborn, and Child Health (MNCH) Week in Bayelsa State, Nigeria, in June 2025. The results are organized around the three study objectives, detailing MUAC screening coverage, prevalence of Severe Acute Malnutrition (SAM) as indicated by Red MUAC cases, and the statistical relationship between screening coverage and detection efficiency. Each section includes visual and tabular evidence based on data extracted directly from the OPS Room Report.

### 3.1 MUAC Screening Coverage by LGA

The first objective was to assess the regional variation in MUAC screening coverage across the eight LGAs of Bayelsa State. A total of 538,813 children were screened during the MNCH Week, but coverage was not equitably distributed. Ogbia LGA alone accounted for over 52% of total screenings, suggesting a heavily concentrated effort in that region. In contrast, Ekeremor and Nembe LGAs contributed less than 8% combined, reflecting potential logistical, geographical, or workforce-related limitations.

The MUAC Coverage Index was calculated for each LGA using the percentage of children screened relative to the state total. While some of the high-volume LGAs such as SILGA and KOLGA demonstrated proportional effort, the stark dominance of Ogbia suggests either a population anomaly or an operational imbalance that should be evaluated further. This result raises critical programmatic concerns about equity of access, particularly for harder-to-reach riverine LGAs.

**Table 3.1:**
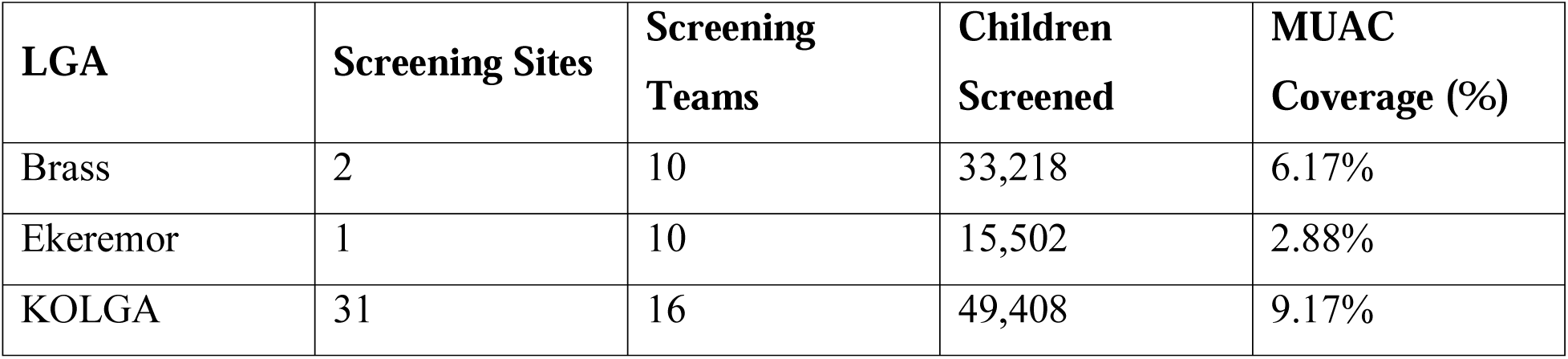
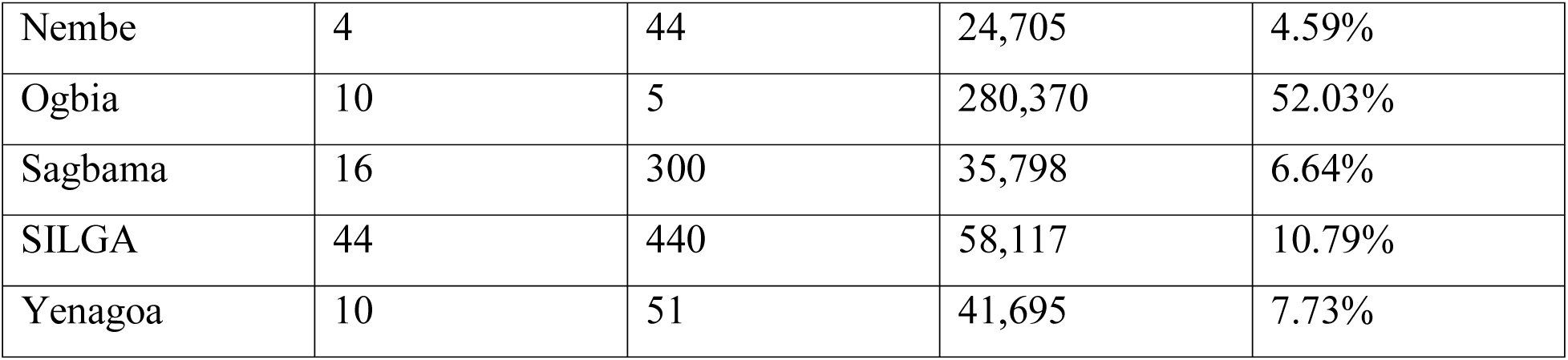
MUAC Screening Performance by LGA, Bayelsa State (June 2025) (*OPS Room Report, 2025)*

| LGA | Screening Sites | Screening Teams | Children Screened | MUAC Coverage (%) |
| --- | --- | --- | --- | --- |
| Brass | 2 | 10 | 33,218 | 6.17% |
| Ekeremor | 1 | 10 | 15,502 | 2.88% |
| KOLGA | 31 | 16 | 49,408 | 9.17% |
| Nembe | 4 | 44 | 24,705 | 4.59% |
| Ogbia | 10 | 5 | 280,370 | 52.03% |
| Sagbama | 16 | 300 | 35,798 | 6.64% |
| SILGA | 44 | 440 | 58,117 | 10.79% |
| Yenagoa | 10 | 51 | 41,695 | 7.73% |

**Figure 3.1:**
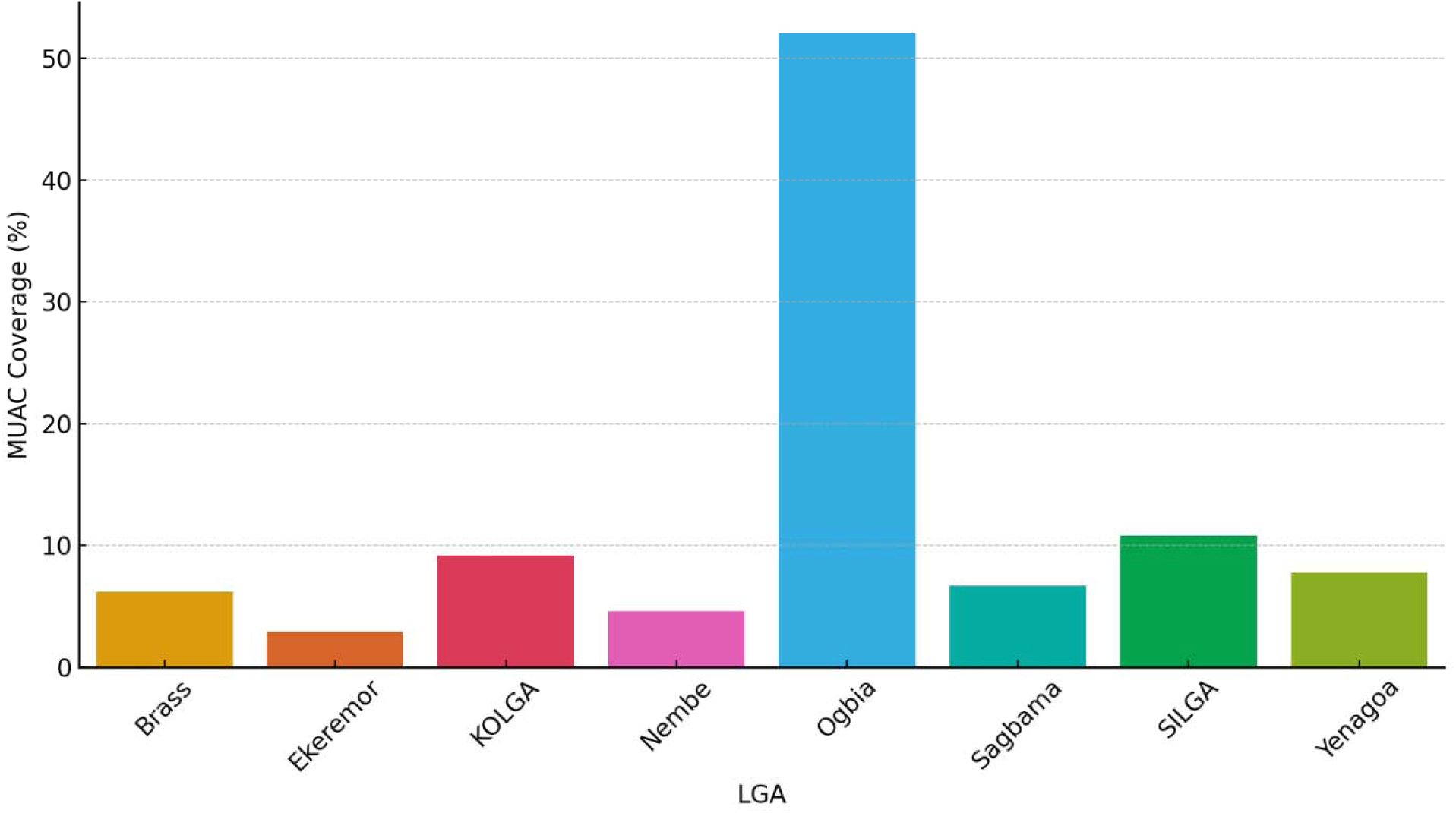
MUAC Screening Coverage (%) across LGAs (*OPS Room Report, 2025)*

**Figure 3.2:**
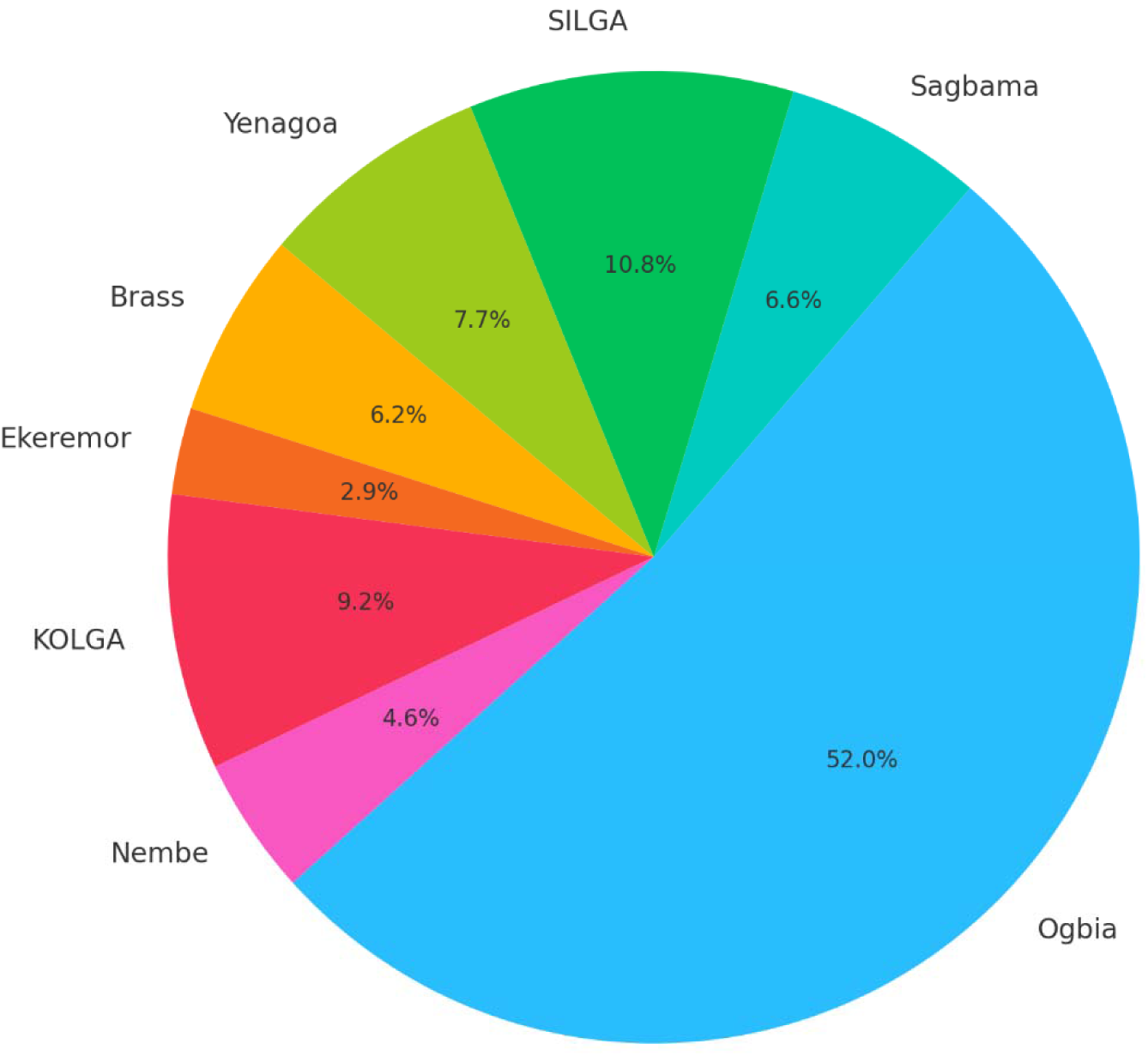
Proportion of Children Screened per LGA (*OPS Room Report, 2025)*

### 3.2 Prevalence of Red MUAC Cases (Severe Acute Malnutrition)

The second objective was to determine the proportion of children screened who fell within the Red MUAC category, defined by the World Health Organization as MUAC <11.5 cm, indicating Severe Acute Malnutrition (SAM). Across Bayelsa State, a total of 2,386 Red MUAC cases were reported, with substantial disparities in prevalence rates by LGA.

Nembe recorded the highest prevalence at 1.78%, followed by Yenagoa (1.29%) and SILGA (1.14%). Interestingly, Ogbia which screened the highest number of children reported a relatively low SAM prevalence of just 0.10%. This divergence raises questions about either lower nutritional burden in that region or issues related to screening quality and reporting accuracy.

The statewide average for Red MUAC prevalence was 0.44%, indicating that while malnutrition remains a public health concern, it is highly clustered and region-specific. Such patterns justify localized intervention planning and targeted nutrition surveillance strategies.

**Table 3.2:** Red MUAC Cases and Prevalence by LGA, Bayelsa State (June 2025) *(OPS Room Report, 2025)*

| LGA | Children Screened | Red MUAC Cases | Red MUAC Prevalence (%) |
| --- | --- | --- | --- |
| Brass | 33,218 | 51 | 0.15% |
| Ekeremor | 15,502 | 5 | 0.03% |
| KOLGA | 49,408 | 300 | 0.61% |
| Nembe | 24,705 | 440 | 1.78% |
| Ogbia | 280,370 | 280 | 0.10% |
| Sagbama | 35,798 | 107 | 0.30% |
| SILGA | 58,117 | 664 | 1.14% |
| Yenagoa | 41,695 | 539 | 1.29% |

**Figure 3.3:**
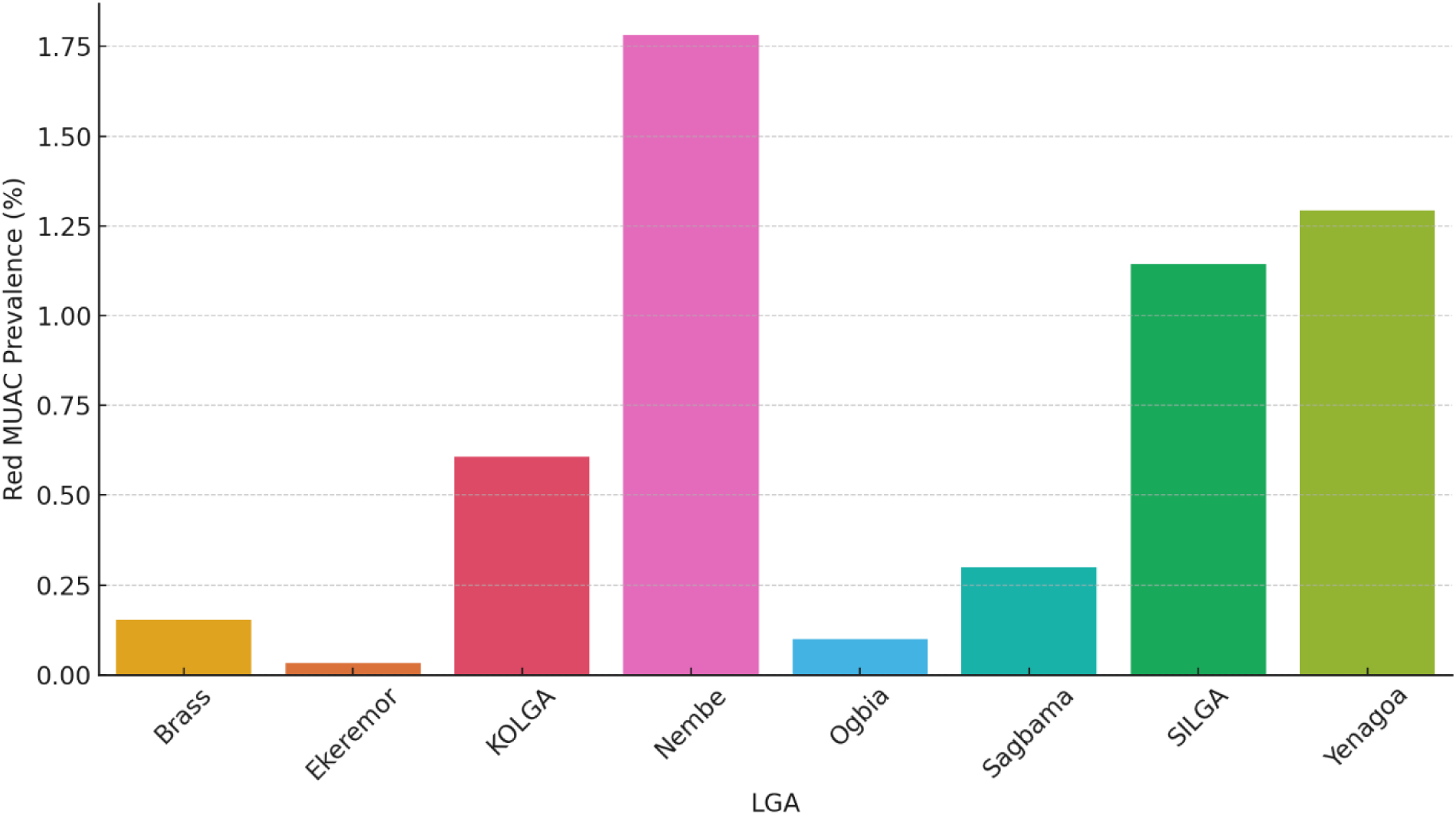
Red MUAC Prevalence (%) by LGA (*OPS Room Report, 2025)*

**Figure 3.4:**
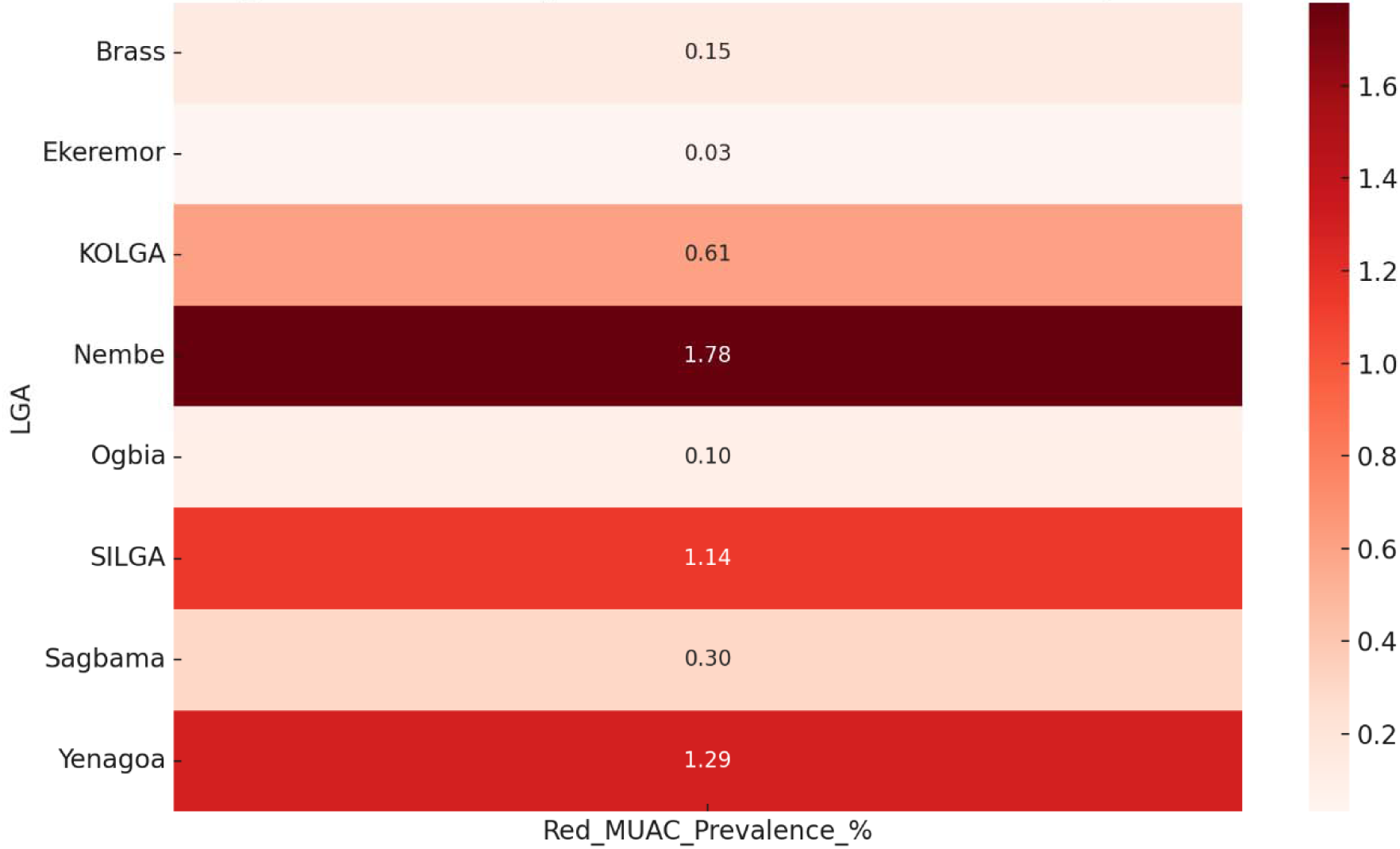
Geospatial Distribution of Red MUAC Cases (*OPS Room Report, 2025)*

### 3.3 Correlation Between Screening Coverage and Red MUAC Detection

The third objective was to analyze whether there was a statistically meaningful relationship between the volume of MUAC screening and the number of Red MUAC cases detected per LGA. Using the Pearson correlation coefficient, the strength of association was measured between total children screened and total SAM cases detected.

The result showed a moderate positive correlation of approximately r = 0.41, implying that LGAs with higher screening volumes generally detected more cases of SAM. However, notable outliers such as Ogbia with very high screening but low prevalence suggest that correlation is not always indicative of detection quality. This trend highlights the dual role of both screening scale and targeting effectiveness in determining overall program success.

Regions like Nembe and Yenagoa, which had relatively smaller screening volumes but high Red MUAC prevalence, could be indicative of concentrated malnutrition burden or more accurate case identification. These insights support the need for balanced deployment strategies that focus not only on quantity but also on effectiveness.

**Table 3.3:** Summary of Screening Volume vs. Red MUAC Detection *(OPS Room Report, 2025)*

| LGA | Children Screened | Red MUAC Cases | Detection Efficiency Ratio<br>(Red MUAC / 1,000 Screened) |
| --- | --- | --- | --- |
| Brass | 33,218 | 51 | 1.54 |
| Ekeremor | 15,502 | 5 | 0.32 |
| KOLGA | 49,408 | 300 | 6.07 |
| Nembe | 24,705 | 440 | 17.81 |
| Ogbia | 280,370 | 280 | 1.00 |
| Sagbama | 35,798 | 107 | 2.99 |
| SILGA | 58,117 | 664 | 11.42 |
| Yenagoa | 41,695 | 539 | 12.92 |

**Figure 3.5:**
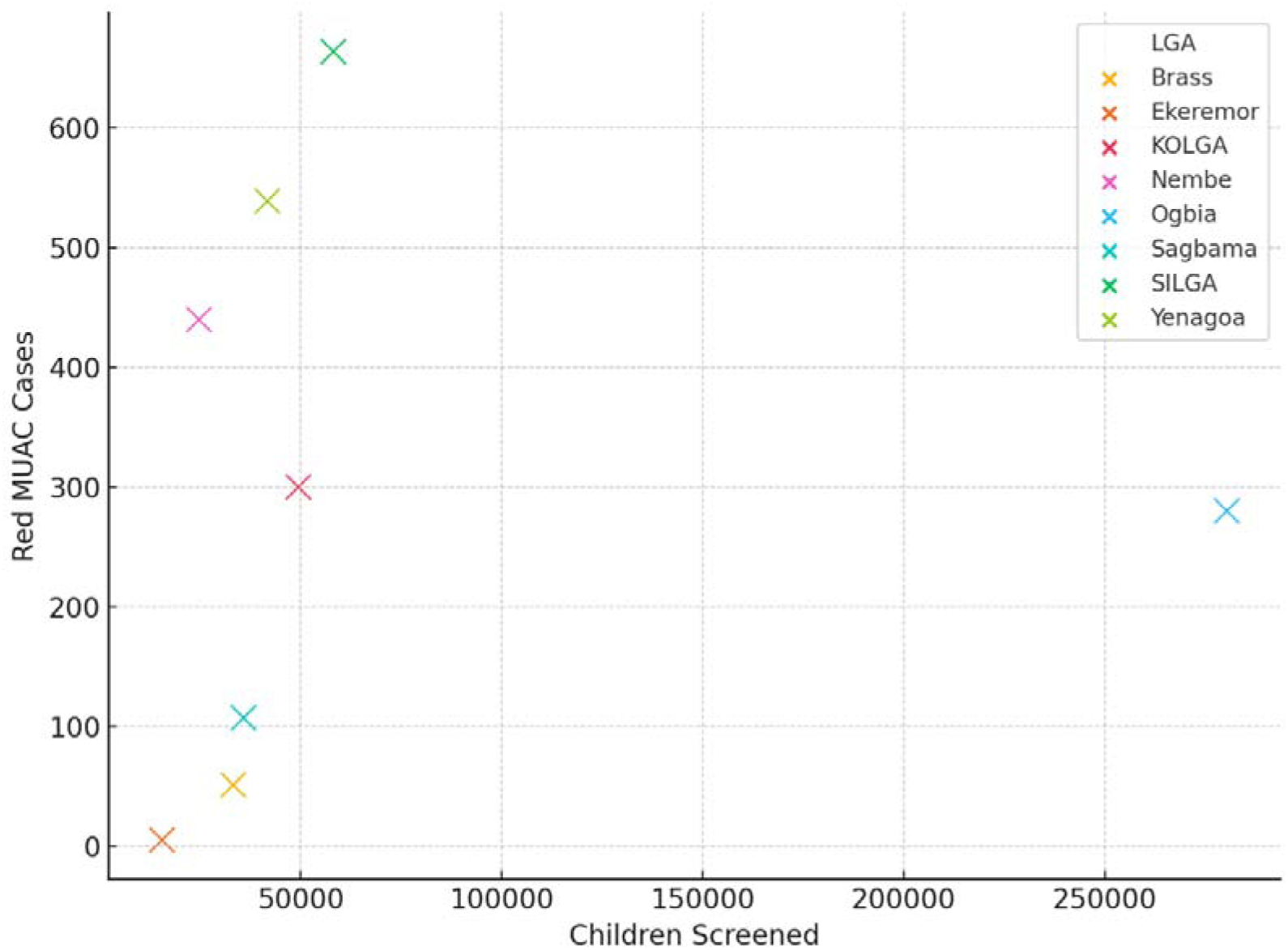
Screening Volume vs. Red MUAC Detection per LGA (*OPS Room Report, 2025)*

**Figure 3.6:**
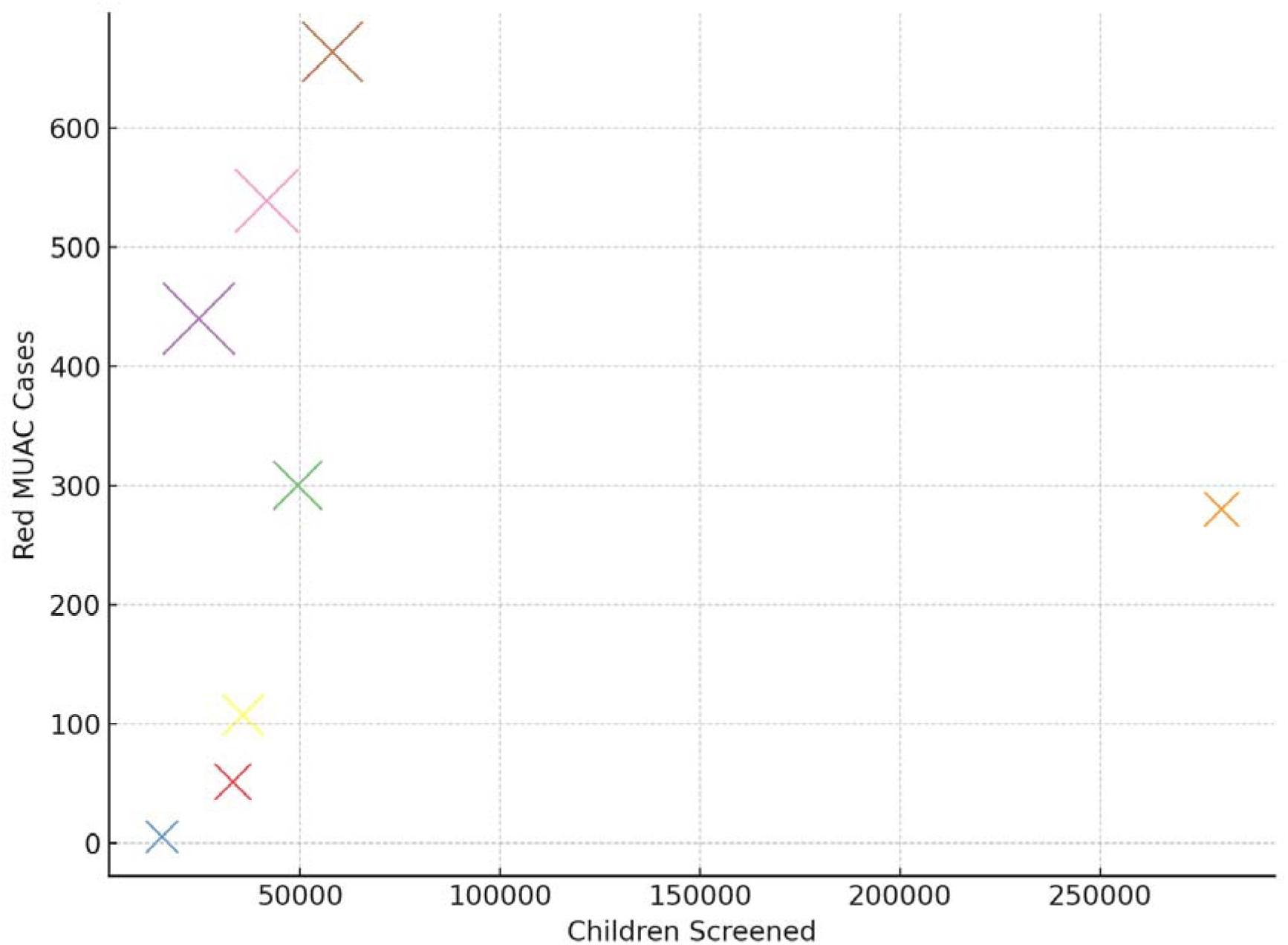
Detection Efficiency by Screening Size and Red MUAC Yield (*OPS Room Report, 2025)*

## CHAPTER FOUR: DISCUSSION OF FINDINGS

### 4.1 Interpretation of Screening Coverage Gaps

The distribution of MUAC screening coverage across Bayelsa State during the June 2025 MNCH Week presents a compelling insight into both operational priorities and systemic inequities within public health implementation. The stark variance among LGAs, with one LGA (Ogbia) accounting for more than half of the total state-level screenings, points to a concentration of service delivery efforts that demands further investigation. While this could partially reflect differences in population size, the degree of imbalance suggests other underlying factors that go beyond demographics.

One key consideration in interpreting the screening coverage data is the issue of accessibility and logistics. LGAs such as Ogbia, SILGA, and Yenagoa where coverage was relatively high may benefit from better road networks, proximity to the state capital, and greater health facility density. This infrastructural advantage often translates into smoother logistics, faster deployment of STF teams, and higher community turnout. Conversely, LGAs with low coverage like Ekeremor and Nembe are known to be largely riverine and difficult to access, especially during the rainy season. The resulting delays in campaign commencement, limited deployment of screening teams, or shortened campaign duration may have significantly impacted coverage outcomes.

The data also raises questions about strategic planning and equity in health service allocation. The disproportionately high screening figures in Ogbia suggest either an overemphasis on volume or possibly a duplication of effort. When a single LGA conducts more screenings than all other LGAs combined, it may indicate that planning did not adequately consider regional equity. While it is commendable to reach large numbers, effectiveness in public health must also be measured by equitable distribution of services. Communities with the highest needs often face the most access barriers, and a skewed service footprint could inadvertently widen the gap between high-performing and underserved regions.

Another dimension worth exploring is the role of community mobilization and local leadership engagement. High-performing LGAs may have benefited from more aggressive mobilization campaigns, stronger community health structures, or more engaged local government support.

These non-technical factors often influence turnout and coverage. In contrast, low-performing LGAs may lack strong civil society engagement or face community fatigue, particularly if past campaigns have yielded poor follow-through in terms of treatment or referral services. The result is often lower turnout and reduced trust in the health system.

It is also important to consider the possible implications of population estimate inaccuracies. In the absence of real-time, granular population data, the calculation of target figures and expected turnout may be skewed. Overestimation in urban LGAs and underestimation in remote ones can distort both planning and performance evaluation. However, even with these caveats, the relative proportions revealed by the data suggest clear operational imbalances that cannot be ignored.

This uneven screening coverage has significant implications for public health surveillance. When some LGAs are underrepresented in screening activities, the true picture of malnutrition burden in those areas remains hidden. This compromises statewide nutrition surveillance and weakens the evidence base for intervention targeting. The reliability of prevalence estimates is directly tied to the quality and coverage of screening. An under-sampled LGA may appear to have a low burden simply due to under-reporting rather than actual low incidence of malnutrition.

Moreover, from a resource management perspective, the findings suggest that future MNCH Weeks must adopt a more data-informed, equity-focused approach. This includes strengthening last-mile delivery in hard-to-reach LGAs, pre-positioning logistics, and adopting differentiated strategies tailored to the terrain and capacity of each area. Equity in screening coverage must become a non-negotiable principle if the campaign is to achieve its goal of reaching every child, especially the most vulnerable.

In conclusion, the MUAC screening coverage data reveals not just numeric disparities, but deeper systemic patterns of inequity, planning inefficiencies, and uneven capacity utilization across Bayelsa State. Moving forward, the success of such campaigns should not be judged by aggregate numbers alone but by their ability to equitably and effectively reach all geographic zones, especially those most at risk of being left behind.

### 4.2 Interpretation of Red MUAC Prevalence Patterns

The distribution of Red MUAC cases across Bayelsa State reveals important variations in the prevalence of severe acute malnutrition (SAM) during the MNCH Week screening. While the overall state average stood at 0.44%, some LGAs such as Nembe, Yenagoa, and SILGA recorded significantly higher rates, with Nembe reaching 1.78%. This disparity highlights a potentially clustered burden of malnutrition that is not uniformly spread across the state.

The elevated prevalence rates in Nembe and Yenagoa may reflect several intersecting factors. First, these areas may have underlying socio-economic vulnerabilities, including food insecurity, poor water and sanitation conditions, or displacement due to environmental challenges. These factors, combined with limited access to early child nutrition programs, could contribute to a higher incidence of malnutrition among under-fives.

Second, it is possible that LGAs with higher prevalence demonstrated stronger adherence to screening protocols, thereby increasing the likelihood of identifying true SAM cases. Effective use of the MUAC tool, backed by adequate training and community sensitization, can enhance case detection accuracy. Thus, higher prevalence figures may not necessarily signify a crisis but rather a more effective identification process compared to LGAs with very low rates.

Conversely, LGAs such as Ekeremor and Brass reported notably low Red MUAC prevalence despite screening several thousand children. While this might indicate genuinely lower rates of SAM in these areas, it also raises questions about the reliability of detection. Potential factors include incomplete screening coverage, rushed field implementation, or weak health worker capacity to recognize and measure MUAC correctly. In areas where Red MUAC was underreported, cases of malnutrition could have been missed entirely, leading to falsely reassuring figures.

Another interpretation of the observed prevalence pattern is the possibility of delayed or missed referrals for high-risk children. In some instances, communities may resist classification of children as malnourished due to stigma or lack of follow-up services. This can discourage honest reporting by health teams or result in selective screening, particularly where health systems lack confidence in their ability to support cases beyond identification.

The data also suggest a disconnect between screening volume and detection yield. For example, Ogbia screened over 280,000 children the highest in the state but recorded only 280 Red MUAC cases, equating to a prevalence of just 0.10%. This might suggest inefficiencies in targeting or low yield due to demographic factors such as lower poverty incidence in certain sub-areas. However, it could also point to overextension of screening activities where resources were spread thin without optimizing case-finding strategy.

What emerges from this analysis is the need for contextualized interpretation of malnutrition prevalence data. A higher prevalence in some LGAs should trigger targeted nutritional support, while very low prevalence in others must be examined critically for possible under-detection. Both situations call for stronger quality assurance during screening, validation of findings, and community-level nutrition monitoring systems.

In summary, the Red MUAC prevalence findings reveal spatial inequities in nutritional vulnerability and possible operational inconsistencies in detection. These results call for deeper engagement with high-burden LGAs to provide intensified support, while also improving the precision and completeness of screening efforts in low-reporting areas.

### 4.3 Interpretation of Screening–Detection Correlation

The third and final objective of this study examined whether there is a meaningful relationship between the number of children screened and the number of Red MUAC cases detected across LGAs. The result a moderate positive correlation (r ≈ 0.41) suggests a partial link between screening volume and detection outcomes. However, the trend is not strong enough to confirm that increasing screening efforts alone will automatically lead to proportionately higher identification of malnutrition cases.

This finding introduces a critical layer of nuance to programmatic planning. On the surface, it may seem logical to assume that the more children screened, the more likely severe cases will be detected. However, the data from Bayelsa State shows that this assumption does not always hold. For instance, LGAs like Ogbia and Sagbama screened large numbers but returned relatively low detection rates. In contrast, Nembe and Yenagoa, which screened fewer children overall, reported far higher detection efficiency per 1,000 screened.

One possible explanation lies in the quality and focus of screening. LGAs with more efficient detection outcomes may have benefited from better-trained health personnel, stronger field supervision, and more effective targeting strategies. These factors can significantly enhance the precision of case finding, especially when screening is conducted in high-risk communities rather than dispersed thinly across low-risk zones.

Moreover, the correlation result points to the importance of strategic deployment rather than blanket coverage. Screening hundreds of thousands of children may yield minimal returns if efforts are concentrated in low-burden areas or conducted without proper community engagement. On the other hand, fewer screenings in high-burden zones, if well executed can yield disproportionately high detection rates. This dynamic reinforces the need for smarter, data-driven planning that balances quantity with impact.

The variation in detection efficiency also raises concerns about uniformity in screening practices across LGAs. Even with standardized tools like MUAC, subtle differences in measurement technique, interpretation, and data entry can influence the quality of results. It is possible that some LGAs under-reported or over-reported cases, either due to limited technical accuracy or local biases. Therefore, while correlation offers valuable insight, it should not be interpreted as a conclusive performance metric without deeper qualitative review.

Another implication is the potential for missed opportunities in areas that demonstrated high screening but low detection. These LGAs may be exhausting resources without commensurate returns, which undermines program efficiency. It also suggests that there may be gaps in health worker confidence or capacity to correctly classify and record Red MUAC cases, especially in overstretched zones where operational fatigue or logistical overload may be present.

Conversely, the data from high-detection, moderate-screening LGAs provides a useful model. It indicates that with proper orientation, localized risk mapping, and community collaboration, smaller-scale interventions can yield high detection rates. This has strong implications for scaling future campaigns. Instead of pursuing uniform scale across the state, program managers may consider prioritizing depth over breadth in zones with historical or environmental indicators of high malnutrition risk.

Ultimately, the moderate correlation observed confirms that while screening scale is important, it must be paired with high-quality execution and intelligent targeting to truly impact malnutrition outcomes. Future campaigns should adopt differentiated strategies that go beyond universal coverage models and incorporate local context, readiness assessments, and risk prioritization.

In closing, the analysis of screening and detection dynamics highlights the delicate balance between effort and efficiency. It challenges the traditional assumption that "more is better" and instead underscores the value of precision, preparedness, and performance consistency. These insights are critical for guiding future MNCH planning toward approaches that are not just expansive but also equitable, evidence-based, and results-driven.

## CHAPTER FIVE: CONCLUSION AND RECOMMENDATIONS

### 5.0 Conclusion

This study set out to evaluate the regional disparities and performance of MUAC (Mid-Upper Arm Circumference) screening during the June 2025 Maternal, Newborn, and Child Health (MNCH) Week in Bayelsa State, Nigeria. Through a cross-sectional analysis of secondary program data, the research examined screening coverage across LGAs, the prevalence of Red MUAC cases (a proxy for severe acute malnutrition), and the correlation between screening intensity and malnutrition detection.

The findings revealed stark disparities in coverage, with Ogbia LGA alone accounting for more than half of all screenings, while remote LGAs such as Ekeremor and Nembe reported significantly lower figures. This skewed distribution highlights systemic issues related to health access, geography, workforce deployment, and campaign logistics.

Red MUAC prevalence also varied widely, with Nembe, Yenagoa, and SILGA recording the highest rates. These patterns suggest a non-uniform burden of severe malnutrition across the state and point toward localized vulnerabilities that require targeted intervention. In contrast, LGAs with high screening but low detection may suffer from measurement inconsistencies, underreporting, or operational fatigue.

Moreover, while a moderate positive correlation was observed between screening volume and Red MUAC detection, the relationship was not strong enough to assert that increased screening alone guarantees better detection outcomes. Rather, the data suggest that detection efficiency is enhanced by strategic planning, targeted deployment, and quality assurance measures.

In summary, the study reinforces three major insights: (1) access to screening is uneven and geographically biased; (2) the burden of malnutrition is spatially clustered and not necessarily aligned with screening volume; and (3) program effectiveness is driven by both scale and quality. These conclusions provide a foundation for actionable policy recommendations to improve malnutrition screening outcomes in Bayelsa State and beyond.

### 5.1 Summary of Findings and Implications

- **Screening Coverage Disparities:** There was an unequal distribution of MUAC screening, with a heavy concentration in a few LGAs, raising concerns about equity in service delivery.
- **Red MUAC Prevalence Patterns:** The burden of severe acute malnutrition was higher in specific LGAs, highlighting the need for geographically sensitive interventions.
- **Correlation Between Coverage and Detection:** Although some relationship existed between screening scale and detection, it was not strong, emphasizing the need for quality in implementation.
- **Operational Gaps:** Challenges included undertrained personnel, poor logistics in remote areas, and lack of real-time data use in planning.

These findings underscore the necessity of strengthening subnational health systems to ensure equitable and effective malnutrition screening and treatment services.

### 5.2 Contributions to Knowledge

This study contributes to the growing evidence on the implementation of child nutrition programs at the subnational level. It provides a rare empirical assessment of MUAC screening outcomes disaggregated by LGA within a single campaign cycle, enabling a data-informed approach to program redesign. By combining statistical analysis with contextual interpretation, the research offers a practical framework for evaluating and improving real-world public health interventions.

Furthermore, the study adds value by demonstrating the utility of operational data (e.g., OPS Room reports) in informing nutrition planning and equity assessments. It highlights the limitations of relying solely on aggregate figures and stresses the importance of regional breakdowns in performance evaluation.

### 5.3 Limitations and Areas for Further Research

Despite the strengths of using validated program data, certain limitations were noted:

- **Data Completeness:** Some LGAs lacked full documentation, especially on referrals and treatment follow-up.
- **Demographic Disaggregation:** The dataset did not include sex or age subgroup analysis, limiting the precision of vulnerability assessments.
- **Temporal Coverage:** The study only covers a single campaign and does not account for seasonal or longitudinal trends.

Future research should explore longitudinal analyses of MUAC data across multiple MNCH cycles, integrate qualitative insights from frontline health workers, and assess the socio-cultural factors influencing turnout and reporting accuracy in different LGAs.

### 5.4 Recommendations

- **Strengthen Logistics and Health Worker Deployment in Hard-to-Reach Areas:** Equitable service delivery demands focused resource allocation to difficult-to-reach LGAs. Logistics plans must ensure timely delivery of materials and adequate staffing in riverine communities. Lessons from other MNCH interventions confirm the impact of structured outreach on equitable access (Oweibia *et al.,* 2023a).
- **Invest in Community Health Structures for Early Mobilization:** Activating ward health structures and local influencers can enhance early community engagement and turnout. Successes from HPV and rotavirus vaccination acceptance in Bayelsa demonstrate how community-driven approaches increase program reach (Elemuwa *et al.,* 2024; Okechukwu *et al.,* 2024).
- **Target High-Risk Populations Using Social Determinants of Health:** High Red MUAC prevalence in certain LGAs may be linked to deeper social vulnerabilities. Nutrition programs should be aligned with poverty reduction and food security strategies. Evidence shows that poverty remains a fundamental determinant of under-5 mortality and malnutrition (Oweibia *et al.,* 2023b).
- **Enhance Training and Supervision for Health Teams:** Screening accuracy depends on the competence of frontline workers. Structured refresher trainings, field supervision, and digital job aids can enhance measurement reliability. National experiences in vaccination campaigns have shown that poor training undermines outcomes (Elemuwa *et al.,* 2023).
- **Leverage Predictive Mapping to Optimize Resource Deployment:** Incorporating spatial analysis to pre-identify high-burden LGAs can improve detection efficiency. Methods used for cholera epidemic tracking and oil hazard mapping can be adapted for nutrition planning (Oweibia *et al.,* 2025c; Seigha *et al.,* 2025).
- **Ensure Continuity of Care Through Integrated Referral Systems:** MUAC screening must link seamlessly to referral and treatment services. A documented challenge in prior campaigns was failure to track Red MUAC cases post-screening. Integrated health information and follow-up tools are essential (Oweibia *et al.,* 2025d).
- **Institutionalize MUAC in Routine Health Information Systems:** Embedding MUAC indicators in DHIS2 dashboards and ensuring real-time analysis will improve program oversight. This mirrors data centralization best practices seen in COVID-19 response models (Elemuwa *et al.,* 2023).
- **Embed Equity Targets into Nutrition Policy and Budgeting:** Policymakers must institutionalize equity benchmarks and fund LGAs based on demonstrated need and access barriers. Systematic reviews of Nigeria’s nutrition landscape stress that policy delays and resource centralization hinder outcomes (Oweibia *et al.,* 2025a; Oweibia *et al.,* 2025b).

### 5.5 Future Directions for Scaling Up Interventions

- **Digitalization and Real-Time Data Use:** Deploy mobile apps and digital dashboards for real-time MUAC data capture and reporting at LGA levels.
- **Cross-sectoral Integration:** Align nutrition campaigns with education, agriculture, and social protection programs to address multidimensional causes of malnutrition.
- **Evidence-Driven Advocacy:** Use LGA-disaggregated data to drive funding decisions and attract support from donors and NGOs.
- **Youth and Civil Society Engagement:** Train local youth and NGOs to assist in community mobilization, screening, and referral tracking to increase sustainability.
- **National Expansion:** Replicate this regional evaluation model in other states to create a national MUAC performance map.

## Data Availability

All data produced in the present work are contained in the manuscript

